# Neuro-Symbolic AI for Automated Pathology Quality Measurement

**DOI:** 10.64898/2026.07.22.26358635

**Authors:** Felix Brann, Lisa Tadele, Colleen Skau, Gregary Bocsi, Alex Clarke

**Affiliations:** Pharos Health, San Francisco, CA, USA; College of American Pathologists, Washington, DC, USA; University of Colorado Hospital Authority, Aurora, CO, USA

## Abstract

**Background:** Clinical quality measurement often relies on manual abstraction of medical records, an approach that is costly, burdensome, and often infeasible for measures requiring interpretation of narrative text; these constraints have shaped measure development itself, filtering out clinically important measures that are too difficult to operationalize. We evaluated whether neuro-symbolic artificial intelligence (NSAI), which combines large language model extraction with symbolic reasoning, could reliably abstract complex quality measures from narrative pathology reports.

**Methods:** The NSAI system decomposes each measure into atomic questions and is aligned to real-world reports through case-based refinement, an iterative human-in-the-loop process. Using 2,000 independently double-abstracted reports, we compared NSAI-based abstraction against trained human abstractors across four pathology quality measures established by the College of American Pathologists.

**Results:** The NSAI system’s agreement with the adjudicated gold standard (Cohen’s *κ* = 0.95) matched or modestly exceeded that of the trained human abstractors measured against the same standard (*κ* = 0.92), with particularly strong performance on Gastrointestinal Metaplasia (CAP 43). In component analyses, case-based refinement drove the largest accuracy gains (up to Δ*κ* = +0.25), whereas architectural decomposition primarily reduced performance variance across language-model backends more than tenfold, a property essential for clinical deployment.

**Conclusions:** These findings suggest that automated abstraction could enable census-level quality measurement, reduce reporting burden, and expand the range of clinically meaningful measures that can be operationalized from narrative clinical documentation.

**Plain-Language Summary:** Checking whether cancer pathology reports meet quality-of-care standards usually requires trained staff to read each report by hand, which is slow and costly. We tested an artificial-intelligence system that combines language models with rule-based logic to do this automatically, and found that it agreed with an expert-reviewed answer key as well as, or slightly better than, trained human reviewers, while producing consistent results across several different underlying AI models. Such a system could let health organizations monitor care quality across every patient record rather than a small sample, and measure aspects of care that are currently too labor-intensive to track.

## 1 Introduction

Clinical quality measures have always been shaped by a tension: the measures that drive the most meaningful improvement are not always operationally feasible. A measure requiring nuanced interpretation of narrative text, contextual reasoning across report sections, or clinical judgment calls that resist simple rules may be clinically important yet operationally infeasible [1]. This constraint has bifurcated measurement into two broad categories. One comprises complex, clinically relevant measures that capture a patient’s disease state, progress, and outcomes but require abstraction of medical records to monitor patterns of care. The other comprises more limited measures that rely on structured discrete data fields and can obscure nuances of care. Manual abstraction is burdensome, costly, and prone to bias if not collected by a neutral third party [2], yet critical quality-relevant information consistently resides in unstructured text [3, 4]. The Centers for Medicare and Medicaid Services (CMS) has accordingly moved away from claims-based measures, which do not capture the clinical information needed to improve patient outcomes. Without precise methods to extract information from narrative documentation, a significant barrier remains to the secondary use of electronic clinical data for public reporting, research, and policy [5].

Pathology reporting illustrates these constraints with particular clarity. These reports contain diagnostic, staging, and prognostic information essential for cancer care, yet embedded in complex narrative structures that resist simple extraction [6]. The College of American Pathologists (CAP) has established Cancer Protocols mandating specific documented elements for each cancer type, with synoptic reporting required for accreditation [7]. While synoptic formats improve completeness and enable some structured data capture [8], substantial quality-relevant information remains in narrative sections, and extraction remains challenging for context-dependent targets [9, 10]. They cannot always capture clinically relevant detail, however, and some measures resist operationalization: the CAP’s squamous cell skin cancer measure was restricted to manual data entry in 2025 because automation proved too difficult, showing how clinically important measures requiring contextual interpretation face implementation barriers [11].

Classical natural language processing (NLP) algorithms have had limited success in pathology quality measure assessment [12], but required extensive maintenance and struggled with the high linguistic variability of pathology [13]. Their brittleness burdens the clinicians and practices generating data as well as end users: an existing data set cannot easily be queried for new concepts without training a new algorithm.

Large language models (LLMs) suggested a potential solution. Models like GPT-4 understand clinical text without requiring explicit rules for every linguistic variation, achieving approximately 89% accuracy extracting structured data from lung cancer pathology reports [14, 15, 16]. Yet such figures can overstate real-world reliability, and recent agent-based systems concluded that LLM abstraction is best used as a human-supervised screening aid rather than for autonomous abstraction [17]. LLMs also introduce their own limitations: unreliable, high-variance output, opaque reasoning, reliance on “common sense” rather than precise measure specifications, and fabricated medical information in 25% to 50% of outputs on clinical research tasks [18, 19, 20, 21]. They also lack the traceability needed to pinpoint and act on specific quality gaps with confidence [22]. The need for rigorous validation of AI-extracted clinical data is increasingly recognized; frameworks such as VALID establish structured approaches to assess the accuracy, reliability, and auditability of LLM- and machine learning–derived information from electronic health records[23]. A naive application of LLMs to quality measurement risks replacing one set of problems with another.

Neuro-symbolic artificial intelligence (NSAI) offers a path forward, integrating neural LLMs that parse variable, context-dependent text with symbolic reasoning that enforces explicit rules and provides auditable decision traces [24, 25, 26, 27]. An LLM interprets isolated concepts, such as a tumor’s margin status, while a symbolic structure combines extracted facts into deterministic, auditable outputs [18, 28, 29]. Decomposing a measure into atomic questions constrains each query to a focused extraction task, improving reliability over assessing the whole measure at once and yielding interpretable intermediate results [15, 18, 30]. Because official specifications rarely anticipate the language of real reports, we pair this architecture with case-based refinement, an iterative human-in-the-loop process that adapts both prompts and system structure to observed errors, aligning measure intent with real documentation. Early hybrid systems are promising: Prenosil et al. achieved physician-level accuracy on radiology reports with complete reasoning chains [18], and the Neuro-Symbolic System for Cancer improved oncologic entity extraction by 33% [31]. If such approaches abstract narrative text reliably, interpretably, and consistently, the constraints that have historically filtered measure development may dissolve.

In this paper we validate this methodology through a study of four pathology quality measures established by the College of American Pathologists. Using 2,000 double-abstracted pathology reports, we compared NSAI-based abstraction to trained human abstractors. We find that the NSAI system’s agreement with adjudicated gold standards exceeds the inter-rater reliability of trained abstractors, with substantially lower performance variance across model backends than single-pass approaches.

## 2 Methods

### 2.1 Study Design

We conducted a validation study comparing NSAI-based abstraction of pathology quality measures with manual abstraction by trained human reviewers (Figures 1–2). The primary outcome was abstraction accuracy, defined as concordance between the automated determination and the human-adjudicated gold standard, quantified using Cohen’s kappa. Secondary outcomes were (1) AI and human agreement with the gold standard, and human inter-rater reliability prior to adjudication; (2) systematic differences in error patterns between model variants; and (3) cross-model robustness, quantified as performance variance across frontier language-model backends. This study follows the TRIPOD-LLM reporting guideline; a completed checklist is provided in the Supplementary Appendix.

**Figure 1:**
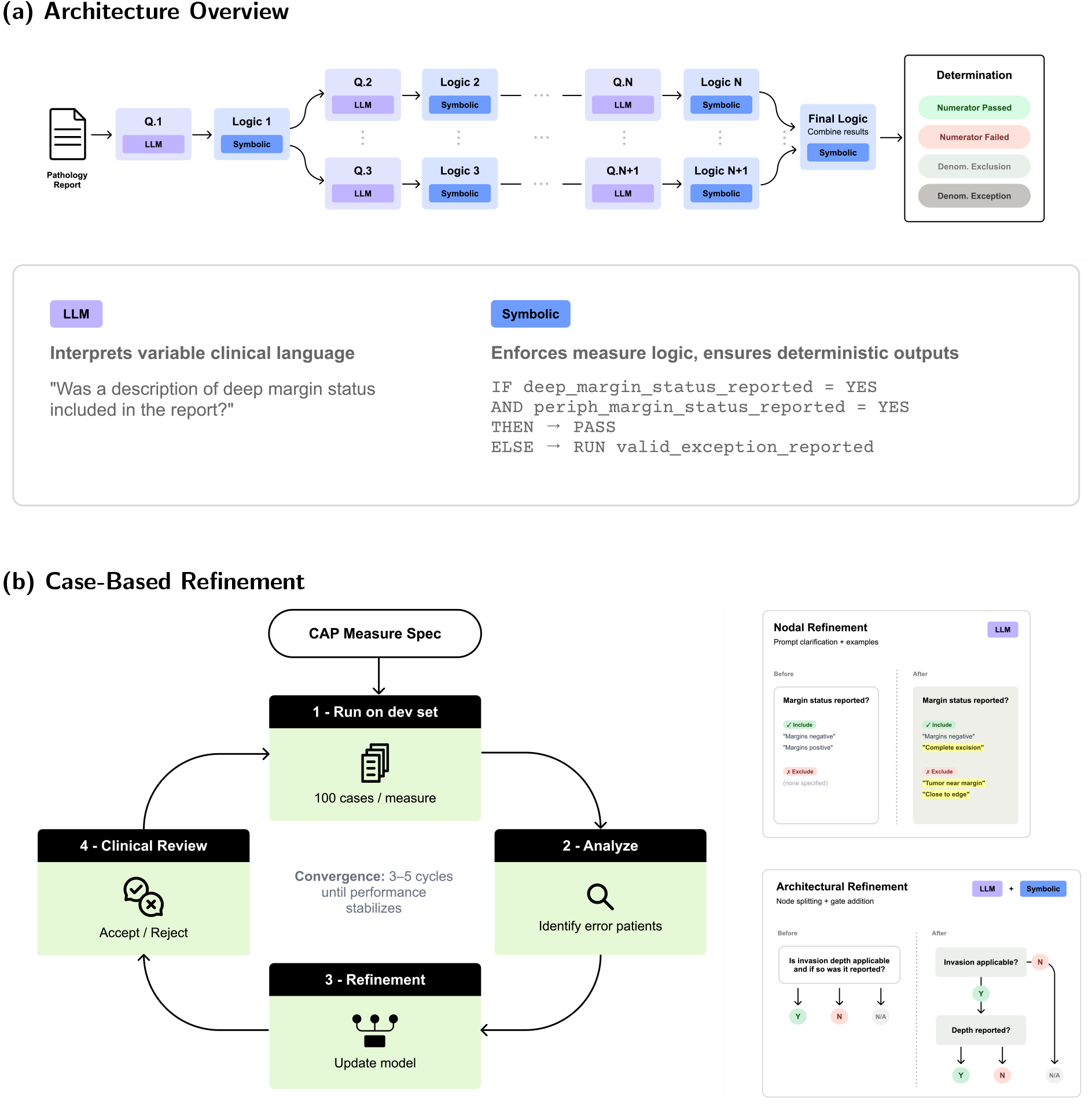
Neuro-symbolic architecture and case-based refinement. (a) The neuro-symbolic AI architecture processes each pathology report through a directed acyclic graph of discrete clinical questions; LLM nodes interpret variable clinical language while symbolic logic nodes enforce measure-specific rules and propagate intermediate results to a final symbolic combiner that produces one of four determinations (Numerator Passed, Numerator Failed, Denominator Exclusion, or Denominator Exception). The lower sub-panel details the two node types. (b) Case-based refinement iteratively aligns the NSAI system with real-world documentation over three to five cycles on the development set: a clinical informaticist reviews errors and accepts or rejects agentic suggestions, which take the form of nodal refinement (prompt clarification and positive/negative examples) or architectural refinement (node splitting and gate addition).

**Figure 2:**
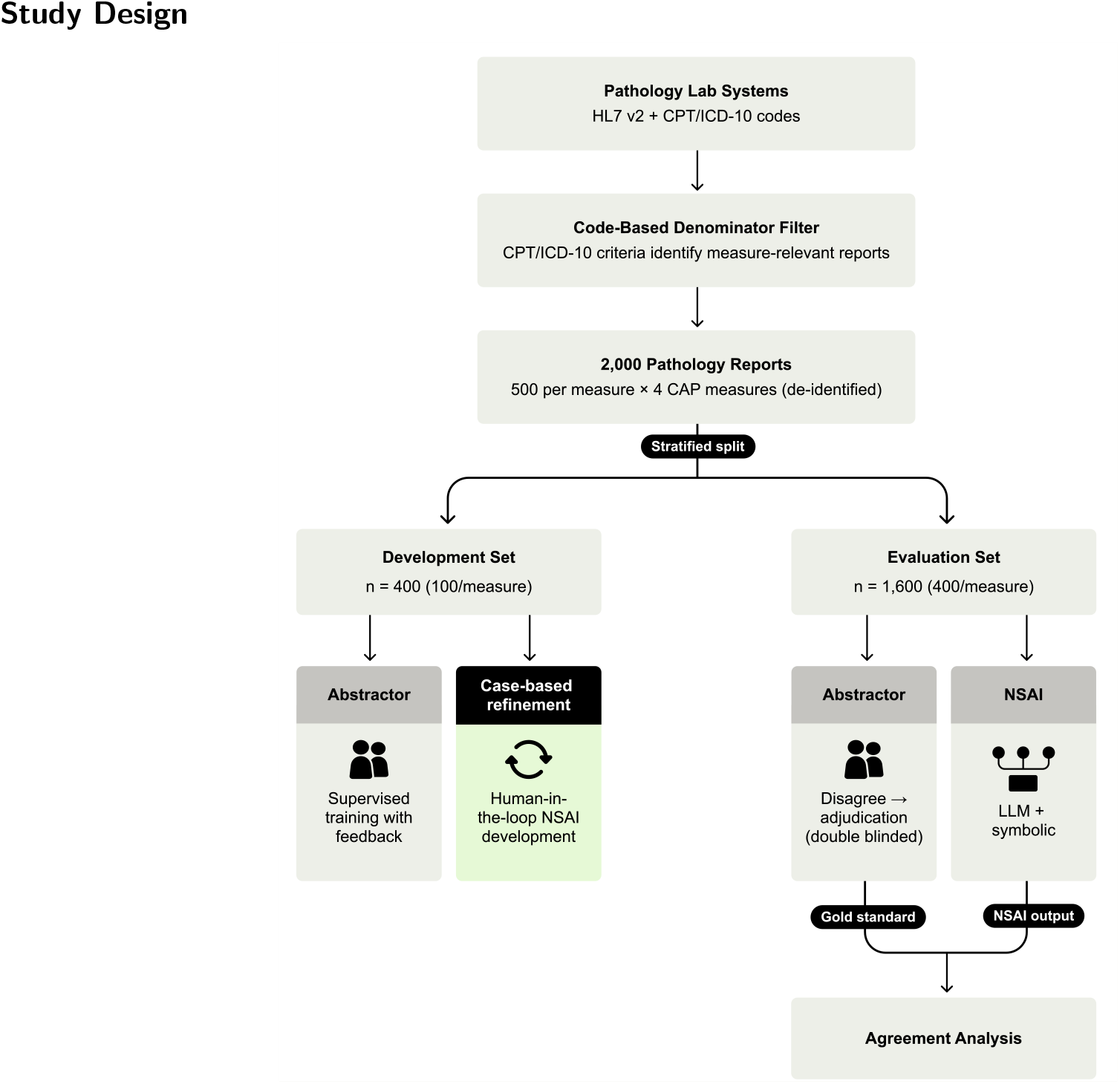
Study design and dataset. Reports were filtered to measure-relevant cases, yielding 2,000 de-identified pathology reports (500 per measure) split into a development set (n = 400) used for abstractor training and NSAI refinement, and a held-out evaluation set (n = 1,600) on which human abstractors performed independent double-blinded abstraction, with disagreements resolved by senior adjudication, and NSAI outputs were compared against the resulting gold standard.

### 2.2 Quality Measures

We evaluated four College of American Pathologists (CAP) pathology quality measures spanning two reporting frameworks, MIPS Clinical Quality Measures and the CAP Qualified Clinical Data Registry: Barrett’s Esophagus (Quality ID #249), Melanoma Reporting (Quality ID #397), Basal Cell Skin Cancer (CAP 41), and Gastrointestinal Metaplasia (CAP 43). They span distinct abstraction challenges, from single-element extraction with conditional grading (Barrett’s Esophagus) through specimen-dependent applicability determination (Basal Cell Carcinoma, Gastrointestinal Metaplasia) to multi-parameter synthesis across report sections (Melanoma); automated extraction of CAP 41 had not previously been feasible with rule-based systems. Full measure specifications are provided in the Supplementary Appendix.

For each report, both AI and human abstractors assigned one of four labels: *numerator passed* (the required quality action was performed and all elements documented), *numerator failed* (required elements absent with no documented reason), *denominator exception* (removed from the denominator only when numerator criteria are not met, reflecting a clinically appropriate deviation), or *denominator exclusion* (removed entirely because the case was never a valid candidate for the measure).

### 2.3 Dataset

Pathology reports, dated between December 2023 and September 2025, were obtained from a US pathology practice. Report components and paired procedure and diagnosis codes (CPT and ICD-10) were received in HL7 v2 format, mapped to a local database using the Fast Healthcare Interoperability Resources (FHIR) data model, and filtered to measure-relevant cases using CPT/ICD-10 denominator criteria. We randomly selected 500 reports per measure (2,000 total), assembled each into a contiguous text document, and screened them manually for completeness; AI and human abstraction used identical formats. The use of these de-identified reports for secondary research was determined exempt by the WCG Institutional Review Board under 45 CFR 46.104(d)(4) (Protocol 20251925), which did not require informed consent. Within each measure, reports were split randomly into a development set (100 per measure; 400 total), used for NSAI refinement and abstractor training, and a held-out evaluation set (400 per measure; 1,600 total) used only for final assessment.

All 2,000 reports underwent independent double-blinded abstraction by two trained clinical abstractors (backgrounds in health information management or nursing) supervised by senior registered nurses; training procedures are detailed in the Supplementary Appendix. Disagreements were resolved by an independent senior reviewer, and the shared result (agreements) or adjudicated result (disagreements) formed the gold standard, with individual determinations retained to characterize inter-rater variability. All human abstraction and adjudication were performed independently of the system’s developers and without access to its outputs, so the gold standard was defined separately from the automated pipeline.

### 2.4 Neuro-Symbolic AI Architecture and Development

The NSAI system pairs LLMs that parse variable clinical language with symbolic reasoning that enforces measure logic [26, 32, 33, 34]. Each measure was formalized as a directed acyclic graph of discrete clinical questions in which LLM nodes perform atomic extractions and symbolic nodes combine them under explicit rules to yield one of the four determinations (Figure 1a). Gemini 2.5 Pro was selected for node inference after evaluating frontier models on the development set. Architecture and node design are detailed in the Supplementary Appendix.

### 2.5 Measure Encoding and Case-Based Refinement

Because official measure specifications do not anticipate the linguistic patterns and edge cases of real reports [35, 36], we operationalized each measure through a human-in-the-loop process, case-based refinement (Figure 1b). Initial measure representations, derived from CAP abstraction guidance via an agentic semi-automated process [37] and reviewed by a clinical informaticist, were iteratively refined on the 400-report development set: system errors were analyzed and the agentic system proposed nodal changes (query wording, positive and negative examples) or architectural changes (node splitting, gate addition), which the informaticist accepted, rejected, or revised, typically over three to five cycles per measure. The held-out evaluation set was never accessed during refinement. Full refinement procedures are provided in the Supplementary Appendix.

### 2.6 Component Comparison Models

To isolate each component’s contribution, we compared the full NSAI system against two single-pass baselines: a *monolithic* model, given the refined measure logic as brief sequential prompt instructions, and a *direct* model, given only the raw CAP specification. The NSAI-versus-monolithic comparison isolates architectural decomposition; the monolithic-versus-direct comparison isolates case-based refinement. Full baseline definitions appear in the Supplementary Appendix.

### 2.7 Statistical Analysis

The analysis pipeline is summarized in Figure 2. Agreement with the adjudicated gold standard was assessed using Cohen’s kappa (*κ*) with 95% bootstrap confidence intervals (2,000 iterations) [38], interpreted via Landis and Koch criteria [39]; differences in kappa (Δ*κ*) were tested by paired bootstrap resampling with one-tailed p-values. AI performance was compared against each human abstractor’s agreement with the gold standard (using the better-performing abstractor as a conservative benchmark) and against pre-adjudication inter-rater reliability. Cross-model stability across five frontier LLMs (Gemini 2.5 Pro/Flash, GPT-5.2, Claude Opus/Sonnet 4.5) was quantified using Levene’s test [40] and Cohen’s *d* [41]. Benjamini-Hochberg correction controlled the false discovery rate at 5% for per-measure comparisons [42]; pooled analyses were treated as primary. Analyses used Python 3.13 with SciPy 1.14 [43] and scikit-learn 1.5 [44].

## 3 Results

### 3.1 Human Abstractor Performance

Human abstractors demonstrated substantial inter-rater reliability prior to adjudication, with a pooled Cohen’s kappa of *κ* = 0.850 and overall agreement of 90.2% across the 1,600 evaluation cases. Interrater reliability varied by measure complexity: Barrett’s Esophagus (Quality ID #249) achieved the highest agreement (*κ* = 0.845), followed by Melanoma Reporting (Quality ID #397; *κ* = 0.670), Basal Cell Carcinoma (CAP 41; *κ* = 0.640), and Gastrointestinal Metaplasia (CAP 43; *κ* = 0.480). These kappas reflect pre-adjudication agreement and should be interpreted in context, as *κ* is sensitive to class prevalence and does not capture how disagreements resolve after expert adjudication.

The low reliability for CAP 43 is consistent with its known abstraction difficulty. When compared against the adjudicated gold standard, individual human abstractors achieved pooled kappa values of *κ* = 0.915 (Abstractor 1) and *κ* = 0.929 (Abstractor 2), though performance varied substantially by measure. The pooled kappa exceeds most individual measure kappas because kappa is sensitive to class prevalence and cases were combined across measures with different outcome distributions.

### 3.2 NSAI System Performance

The NSAI system achieved a pooled agreement with the gold standard of *κ* = 0.954, exceeding the mean performance of the two human abstractors versus gold (*κ* = 0.922; Δ*κ* = +0.032). At the individual measure level, NSAI underperformed the mean human-versus-gold kappa on Barrett’s Esophagus (*κ* = 0.845; Δ*κ* = −0.078), but exceeded the mean human performance on Basal Cell Carcinoma (*κ* = 0.880; Δ*κ* = +0.064), Gastrointestinal Metaplasia (*κ* = 0.829; Δ*κ* = +0.116), and Melanoma (*κ* = 0.947; Δ*κ* = +0.123). In pooled analysis, NSAI also significantly exceeded the best individual abstractor (Δ*κ* = +0.025, *p* = 0.004; Figure 3).

**Figure 3:**
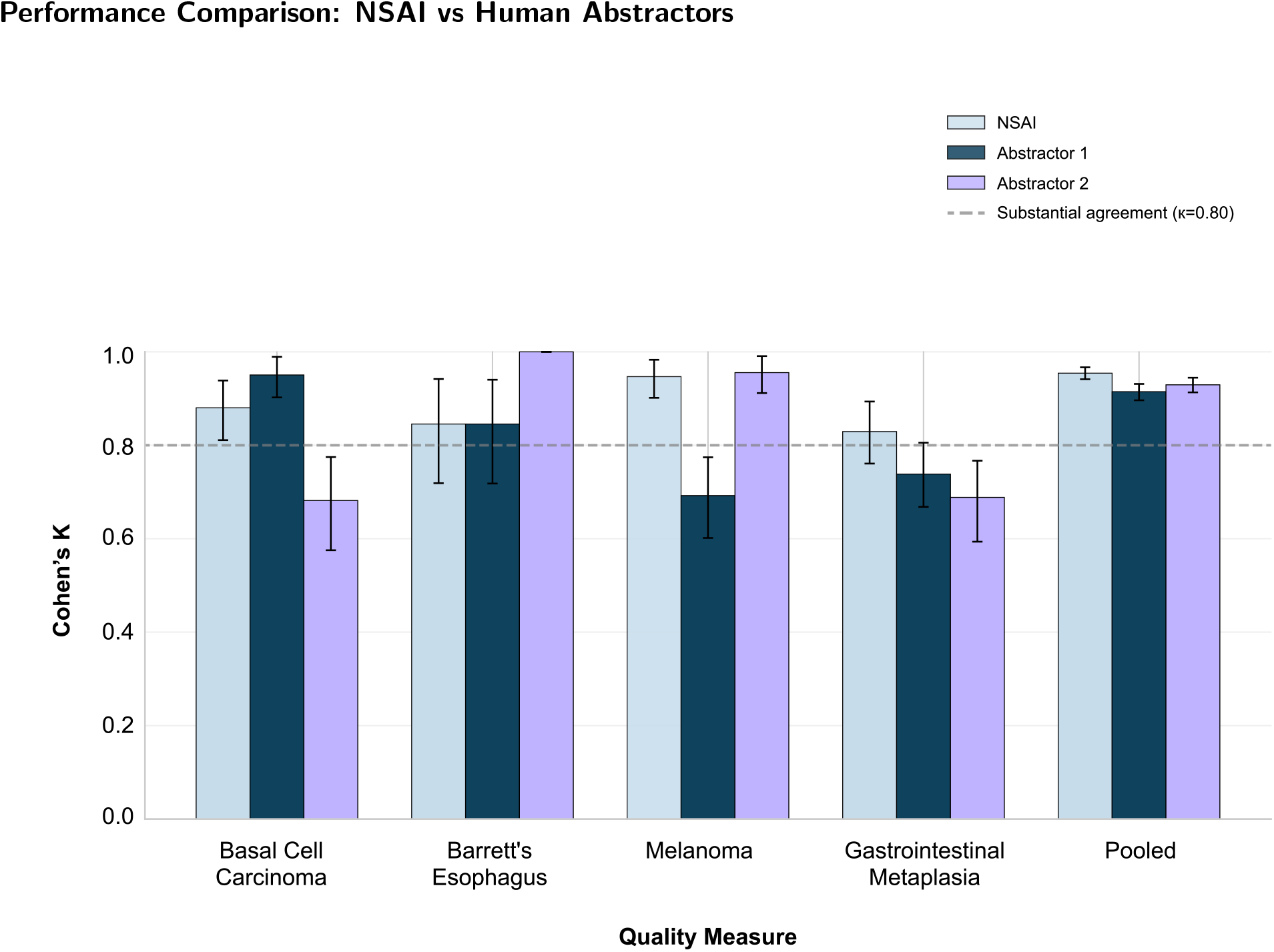
Performance comparison of the neuro-symbolic AI (NSAI) system versus two trained human abstractors across four CAP pathology quality measures and pooled results. Bars show Cohen’s *κ* with 95% bootstrap confidence intervals.

The NSAI system produced 47 errors across 1,600 cases (2.9% error rate). The most common pattern was false negative compliance determination: 11 cases classified as numerator failed when the gold standard was numerator passed, and 8 cases classified as denominator exception when the gold standard was numerator failed. Denominator misclassification accounted for 15 errors (32%). This error type concentrated in Basal Cell Carcinoma, where distinguishing excision from biopsy required contextual interpretation of procedure descriptions. Figure 4 summarizes pooled misclassifications.

**Figure 4:**
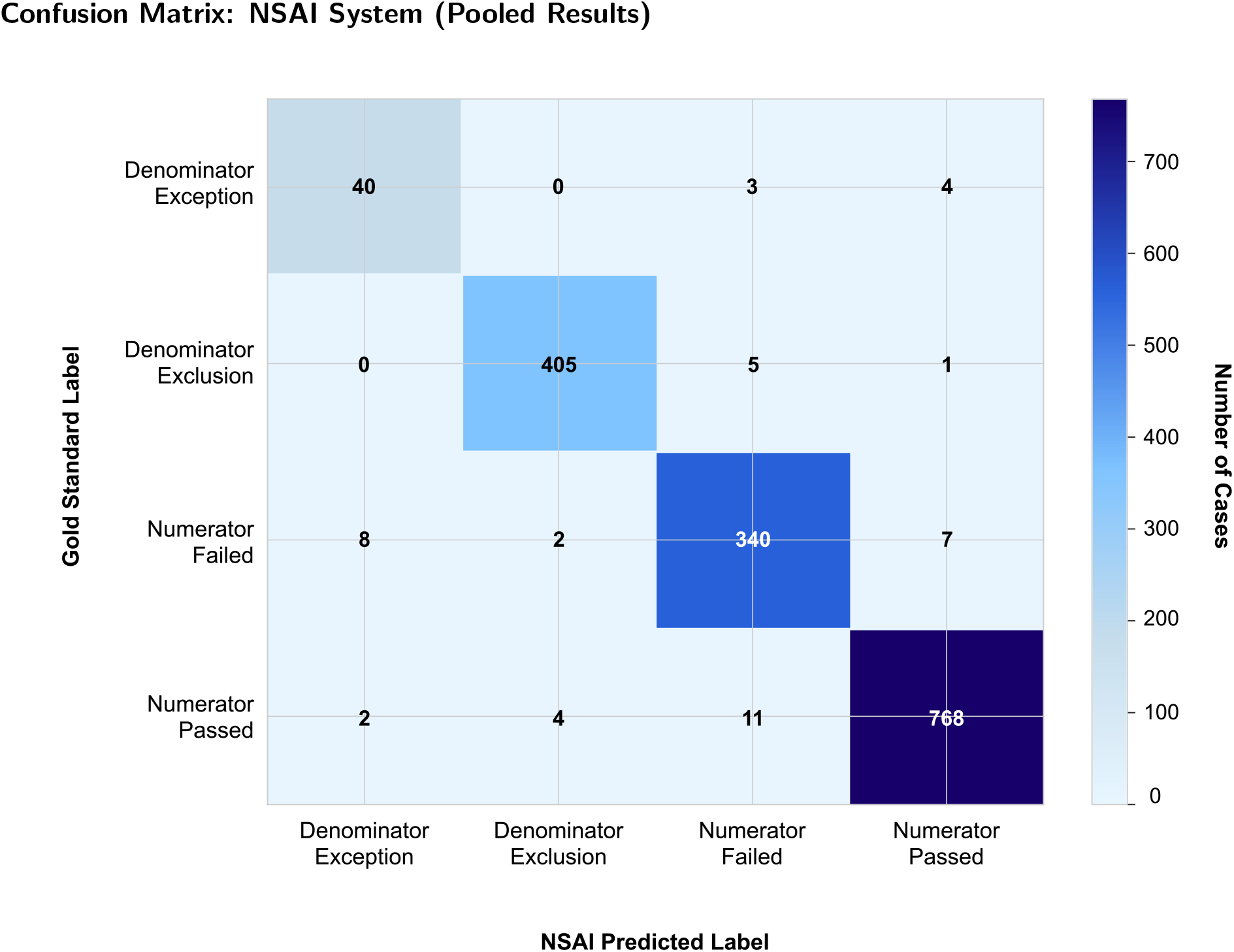
Confusion matrix for the NSAI system on pooled results across all four measures (1,600 evaluation cases). Rows indicate gold-standard labels and columns indicate NSAI-predicted labels; cell values are counts.

### 3.3 Architectural Decomposition

We compared the NSAI architecture with a monolithic variant that used the same refined prompts but executed the task in a single inference pass. Inspection of the monolithic model’s reasoning traces suggested that it often reconstructed the symbolic reasoning hardcoded in the NSAI system, but with substantial model-dependent variability. On a single backbone (Gemini 2.5 Pro), decomposition modestly improved agreement over monolithic prompting (Δ*κ* = 0.020); its larger benefit emerged across models. Across five frontier models the NSAI architecture achieved mean *κ* = 0.827 (SD = 0.077), compared with mean *κ* = 0.634 (SD = 0.254) for monolithic prompting. The NSAI system exhibited 10.8 *×* lower variance than the monolithic alternative (Levene’s *W* = 9.87, *p* = 0.003; Cohen’s *d* = 1.03; Figure 5). No NSAI model-measure combination fell below substantial agreement thresholds, compared with 7 of 20 monolithic combinations.

**Figure 5:**
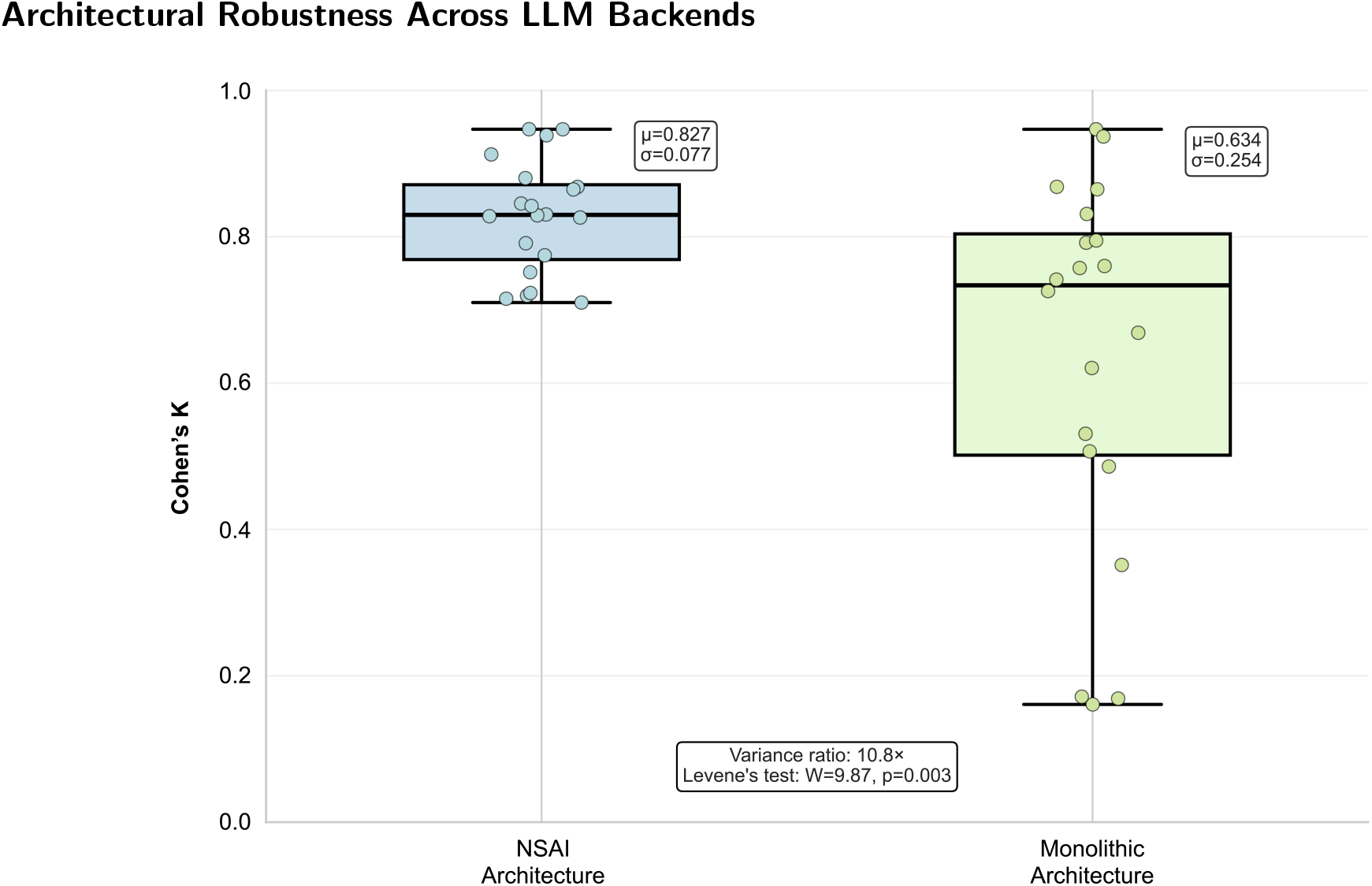
Architectural robustness across five frontier LLM backends. Distributions show Cohen’s *κ* across 20 model–measure combinations (5 models *×* 4 measures) for the NSAI architecture versus a monolithic single-pass prompting baseline.

### 3.4 Case-based Refinement

A second comparison isolated the contribution of case-based refinement by comparing a monolithic prompt that encoded the refined, stepwise question specifications against a direct baseline that used the raw CAP measure specifications verbatim. Although refinement was developed in the NSAI setting, we evaluated it within the monolithic single-pass setup to enable a like-for-like comparison. After Benjamini-Hochberg correction, three measures showed significant improvement: Basal Cell Carcinoma (Δ*κ* = +0.246, *q <* 0.001), Melanoma (Δ*κ* = +0.193, *q <* 0.001), and Gastrointestinal Metaplasia (Δ*κ* = +0.103, *q <* 0.001). Barrett’s Esophagus showed no significant difference (Δ*κ* = −0.025, *q* = 0.68), likely reflecting a ceiling effect where both approaches achieved high performance.

Error analysis revealed that refinement most improved denominator classification: 43 cases where the direct baseline incorrectly assigned denominator exclusion to denominator exception were corrected by the refined prompts. These are among the largest accuracy gains observed in the study, indicating that refinement acts chiefly by resolving the ambiguous applicability logic (numerator, denominator, and exception rules) that official specifications leave underspecified.

## 4 Discussion

U.S. physician practices spend more than $15.4 billion annually on quality measure reporting, much of it on manual chart abstraction, and a single acute-care hospital can devote $5 million and 100,000 person-hours each year to reporting alone [45, 46]. Early clinical evidence indicates that automated abstraction can act on this burden directly: LLM-based abstraction has improved reporting of hospital quality measures [47], and randomized trials have since raised compliance with the CMS SEP-1 sepsis bundle by 13 percentage points [48] and matched or exceeded unaided reviewers when abstracting trial-eligibility criteria from unstructured charts [49]. Our findings suggest a path toward sustainable automation: the NSAI system’s agreement with the adjudicated gold standard matched or exceeded that of the trained human abstractors measured against the same standard (*κ* = 0.95 vs. 0.92). This challenges the conclusion, drawn from agent-based pipelines that degrade sharply from synthetic to real-world reports, that automated pathology abstraction is viable only as a human-supervised screening step [17]: delegating measure logic to symbolic rules rather than the language model alone appears to lift that ceiling.

The system’s performance reflects two complementary mechanisms. Architectural decomposition breaks complex measures into atomic extraction tasks governed by symbolic logic, making decisions more dependable by explicitly encoding intermediate reasoning steps. Case-based refinement is essential because measures often assume domain knowledge and operational conventions not fully specified in writing. In our component analyses these roles proved distinct: case-based refinement produced the largest accuracy gains, while architectural decomposition delivered the cross-model robustness essential for deployment.

The most consequential finding for deployment is the reduced performance variance across language-model backends. A system that performs well with one model but fails with another cannot be reliably deployed, where consistent performance must be guaranteed regardless of vendor relationships or model updates. This stability stems from constraining language models to narrowly-scoped extraction tasks and delegating measure logic to symbolic rules, reducing dependence on any single model’s idiosyncratic reasoning.

The system is intended for abstraction of pathology quality measures from narrative reports to support quality measurement and registry reporting, not to guide individual patient care. Because the symbolic layer records an auditable trace for every determination, the system can run autonomously while remaining open to oversight: when a required element is absent or ambiguous, the responsible extraction step is identifiable and can be routed for expert adjudication. Introducing a new measure requires a clinician or informaticist to perform case-based refinement and validation, as done here; routine operation requires no per-report interaction. We did not evaluate how the system should handle degraded or incomplete inputs in operation, a prerequisite for deployment.

Several limitations temper these findings. Reports came from a single pathology group, so conventions may differ across health systems in ways that limit generalizability. The gold standard was human-adjudicated, so the system may optimize for human interpretation rather than measure developers’ intent. We examined only four measures, and performance may differ for other domains or more complex logic. Finally, we evaluated accuracy in isolation and did not assess computational cost, latency, or workflow integration.

If NSAI systems can reliably abstract complex measures from narrative text, the payoff extends beyond efficiency and reduced clinician burden. Quality measurement could shift from periodic sampling to near-census monitoring, enabling faster feedback and earlier detection of care gaps. More fundamentally, the constraints that have long privileged what is easy to abstract over what is clinically meaningful may loosen, allowing measure sets to reflect clinical priorities rather than data-collection limits.

## Data and Code Availability

The pathology reports analyzed in this study contain protected health information and are not publicly available. The code and measure prompts supporting the findings are available from the corresponding author on reasonable request.

