## Supplementary Appendix for "Neuro-Symbolic AI for Automated Pathology Quality Measurement" (from another version of this preprint: version 1, 2026-07-24)

#### 1 Supplementary Methods

##### 1.1 Quality Measure Specifications

The four CAP quality measures and their required reporting elements were:

- **Barrett's Esophagus (Quality ID #249):** Esophageal biopsy reports documenting Barrett's mucosa must include a statement about dysplasia and, if present, the appropriate grade.
- **Melanoma Reporting (Quality ID #397):** Reports for primary malignant cutaneous melanoma must include pT category, tumor thickness, ulceration status, mitotic rate, peripheral and deep margin status, and presence or absence of microsatellitosis for invasive tumors.
- **Basal Cell Skin Cancer: Complete Reporting (CAP 41):** Final reports for cutaneous basal cell carcinoma excisions must include histologic subtype, margin status, and, when applicable to the specimen, invasion beyond the reticular dermis (or level of invasion for Mohs specimens) and perineural invasion.
- **Complete Reporting of Gastrointestinal Metaplasia (CAP 43):** Reports for gastric specimens with intestinal metaplasia must comment on the presence or absence of *H. pylori*, the presence or absence of dysplasia with grade if present, and, if dysplasia is absent, the completeness of intestinal metaplasia.

##### 1.2 Abstractor Training

Manual abstraction was performed by trained clinical abstractors with backgrounds in health information management or nursing, under the supervision of senior registered nurses serving as project managers and quality assurance leads. All abstractors completed a standardized training program comprising didactic instruction on CAP measure specifications and abstraction guidelines, and supervised practice abstraction with feedback on a set of training cases with known determinations.

##### 1.3 Neuro-Symbolic Architecture and Node Design

We adopted the neuro-symbolic paradigm to reflect how experienced abstractors reason through quality measures: determining what type of specimen is present, identifying which reporting elements apply to that specimen type, and systematically checking whether each required element is documented. This reasoning process was formalized as a directed acyclic graph (DAG) of discrete clinical questions, where nodes represent atomic extraction targets and edges represent logical dependencies between concepts. LLM nodes interpret variable clinical language at each step, while symbolic logic nodes enforce measure-specific rules and propagate intermediate results to a final symbolic combiner that produces one of four determinations. Gemini 2.5 Pro was selected for all node inference after assessment of a collection of frontier models on the development set.

##### 1.4 LLM Inference Configuration

All model inference was performed on January 24 and 25, 2026, using the model vendors' hosted application programming interfaces (Gemini models via Google Vertex AI, GPT models via the OpenAI API, and Claude models via the Anthropic API). Five frontier models were evaluated, identified by the following API model strings: `gemini-2.5-pro`, `gemini-2.5-flash`, `gpt-5.2`, `claude-opus-4-5`, and `claude-sonnet-4-5`; Gemini 2.5 Pro served as the backend for the primary neuro-symbolic system. Because these are closed-weight models served through hosted APIs, their training-data cutoff dates are not publicly disclosed by the vendors; we therefore report the inference dates above as the reference point for model versioning. Structured outputs were obtained through JSON-schema-constrained decoding (Gemini and GPT) or through forced tool invocation (Claude, with a maximum of 2,048 output tokens). Sampling parameters, including temperature, top-p, and the random seed, were left at vendor defaults and were not fixed; model outputs are therefore subject to the nondeterminism inherent to these APIs, and exact regeneration of individual outputs is not guaranteed.

##### 1.5 Case-Based Refinement Procedure

Initial representations of each measure were derived from CAP manual abstraction guidance and encoded as structured components within the NSAI framework using an agentic semi-automated specification process, then reviewed by a clinical informaticist to confirm fidelity to the intended clinical definitions and inclusion criteria. Because static review alone was insufficient to identify all ambiguities and edge cases encountered in real-world documentation, both the NSAI structure and associated prompts were refined using the development set of 400 pathology reports. Refinement proceeded in cycles in which system outputs were evaluated across the full development set, error patterns were automatically identified, and targeted adjustments implemented by the agentic system. Adjustments could be nodal (updating query language or adding positive and negative examples) or architectural (splitting a clinical concept into a new node). These modifications were suggested by the agentic system and then accepted, rejected, or refined by the clinical informaticist, continuing until nodal and overall performance stabilized, typically after three to five cycles per measure. The held-out evaluation set of 1,600 cases was not accessed during refinement.

##### 1.6 Component Comparison Baselines

The direct model represented the naive approach of passing the clinical measure and pathology report to a reasoning LLM and asking it to make a decision on a single inference pass; CAP measures were converted to markdown and added to a prompt alongside a brief description of the possible outcomes. The monolithic model was also a single inference pass with a reasoning-capable LLM, but used in its prompt the full content of the refined NSAI question specifications embedded in brief sequential instructions (e.g., "Step 1: Determine specimen type. Step 2: Assess margin status...") guiding the model through the same extraction logic encoded in the NSAI architecture. This isolated the effect of the explicit neuro-symbolic architecture from inference-time reasoning within a single-pass model.

##### 1.7 Ablation Studies

Two ablation studies isolated the contribution of individual system components. The first ablation compared the full neuro-symbolic architecture against a monolithic variant that used the same refined prompts but replaced modular decomposition with unified end-to-end inference over the complete report text. Both systems used the same LLM backbone (Gemini 2.5 Pro). This comparison isolates the effect of architectural decomposition while controlling for prompt content and model capability. The second ablation compared the monolithic architecture against a direct prompting baseline that presented the raw CAP measure specification without case-based refinement from the development set. Both systems used single-pass inference, isolating the contribution of iterative prompt refinement. For both ablations, the difference in kappa ($\Delta\kappa$) was computed with 95% bootstrap confidence intervals (2,000 iterations); statistical significance was determined by whether the confidence interval excluded zero.

To test whether systems exhibited systematically different error patterns, we applied the Stuart-Maxwell test for marginal homogeneity [1], which generalizes McNemar's test to multi-class outcomes. The Stuart-Maxwell test evaluates whether the marginal distributions of two classification systems differ significantly, detecting systematic biases in error direction even when overall accuracy is similar. Class-level performance was assessed by computing precision, recall, and F1 score for each outcome category (numerator passed, numerator failed, denominator exclusion, denominator exception) to identify whether architectural differences preferentially affected specific outcome types.

##### 1.8 Cross-Model Stability

To assess whether neuro-symbolic architecture benefits generalize beyond a single LLM, we evaluated performance across five frontier models: Gemini 2.5 Pro, Gemini 2.5 Flash, GPT-5.2, Claude Opus 4.5, and Claude Sonnet 4.5. Each model was tested under all three architectural conditions (neuro-symbolic, monolithic, direct) across all four quality measures, yielding 60 model–architecture–measure combinations.

Cross-model stability was quantified using Levene's test for equality of variances [2], comparing the variance in kappa values across models between the neuro-symbolic and monolithic architectures. A significant Levene's test indicates that one architecture produces more consistent performance across different LLM backbones. Effect size was estimated using Cohen's $d$ computed on the distribution of kappa values, with $|d| < 0.2$ interpreted as negligible, $0.2 \leq |d| < 0.5$ as small, $0.5 \leq |d| < 0.8$ as medium, and $|d| \geq 0.8$ as large [3]. We additionally computed the variance ratio (monolithic variance / neuro-symbolic variance) to quantify the relative stability of each architecture.

##### 1.9 Multiple Comparison Correction

To account for multiple comparisons across four quality measures within each ablation study, we computed p-values using a permutation test (2,000 permutations) under the null hypothesis of no difference between systems. Permutation p-values were then corrected using the Benjamini-Hochberg procedure to control the false discovery rate at 5% [4]. Statistical significance was set at $\alpha = 0.05$ (two-sided) for all analyses.

##### 1.10 Error Analysis

Error analysis was performed by examining the pooled confusion matrix and categorizing discordant cases by error type: false positive (AI determined compliant when gold standard was non-compliant), false negative (AI determined non-compliant when gold standard was compliant), and denominator misclassification (AI incorrectly determined measure applicability). For the neuro-symbolic system, errors were further traced to specific extraction steps to identify systematic failures. We additionally computed the rate at which each AI system erred on cases where both human abstractors agreed with the gold standard, providing a measure of errors on "clear" cases versus errors attributable to inherent case ambiguity.

All statistical analyses were conducted in Python 3.13 using SciPy 1.11 [5] and scikit-learn 1.3 [6].

#### 2 Supplementary Results

##### 2.1 Human Abstractor Performance

Human abstractors demonstrated substantial inter-rater reliability prior to adjudication, with a pooled Cohen's kappa of $\kappa = 0.850$ and overall agreement of 90.2% across the 1,600 evaluation cases. Inter-rater reliability varied by measure complexity: Barrett's Esophagus (Quality ID #249) achieved the highest agreement ($\kappa = 0.845$), followed by Melanoma Reporting (Quality ID #397; $\kappa = 0.670$), Basal Cell Carcinoma (CAP 41; $\kappa = 0.640$), and Gastrointestinal Metaplasia (CAP 43; $\kappa = 0.480$). The lower reliability observed for CAP 43 is consistent with its recognized abstraction difficulty, which previously led an effort to automate abstraction of the measure to be halted as infeasible, and validates its inclusion as a challenging test case. Individual abstractor performance against the adjudicated gold standard ranged from $\kappa = 0.682$ to $\kappa = 1.000$ across measure–abstractor combinations, with pooled performance of $\kappa = 0.915$ and $\kappa = 0.929$ for the two abstractors respectively.

##### 2.2 Ablation Study: Architectural Decomposition

Comparison of the neuro-symbolic architecture against a monolithic variant using the same LLM backbone (Gemini 2.5 Pro) revealed a statistically significant benefit of modular decomposition. The neuro-symbolic system achieved a pooled kappa of $\kappa = 0.954$ compared to $\kappa = 0.935$ for the monolithic system, yielding $\Delta\kappa = 0.020$ (95% CI: 0.008–0.031). The Stuart-Maxwell test indicated systematic differences in error patterns between architectures ($\chi^2 = 10.43$, $df = 3$, $p = 0.015$).

At the individual measure level, the neuro-symbolic architecture showed numerical advantages for three measures: Barrett's Esophagus ($\Delta\kappa = 0.104$; 95% CI: 0.005–0.203), CAP 43 ($\Delta\kappa = 0.069$; 95%

CI: 0.015–0.122), and Basal Cell Carcinoma ($\Delta\kappa$ = 0.049; 95% CI: −0.017–0.123). For Melanoma Reporting, both architectures achieved identical performance ($\kappa$ = 0.947), suggesting the measure's structure is amenable to either approach. After Benjamini-Hochberg correction for multiple comparisons, no individual measure comparison remained statistically significant at $q \leq 0.05$, though the pooled effect was robust.

### 3 Supplementary Discussion

#### 3.1 Contributions of Architectural Components

The ablation studies decomposed system performance into contributions from architectural decomposition and case-based refinement. These components operate through different mechanisms and showed different effect magnitudes.

Architectural decomposition, breaking quality measure logic into modular extraction steps rather than requesting end-to-end judgments, produced a modest but statistically significant improvement ($\Delta\kappa = 0.020$). This effect likely reflects reduced cognitive load on the language model: rather than simultaneously parsing report text, interpreting measure specifications, and synthesizing a compliance determination, the modular architecture allows each extraction step to focus on a single well-defined concept. The symbolic reasoning layer then assembles extracted facts according to explicit measure logic, ensuring consistency that end-to-end inference cannot guarantee.

Case-based refinement, iteratively improving prompts using development set errors, produced substantially larger gains ($\Delta\kappa = 0.072$, approximately 3.5× the architectural effect). This finding highlights the importance of domain adaptation: generic prompts based on official measure specifications, while seemingly complete, fail to anticipate the linguistic patterns and edge cases present in real pathology reports. The largest improvements from refinement occurred in denominator classification, where distinguishing applicable from non-applicable specimens required learning institution-specific documentation conventions that no generic specification could anticipate.

These findings suggest that practitioners implementing similar systems should prioritize investment in iterative refinement over architectural complexity. A monolithic architecture with well-tuned prompts outperformed a neuro-symbolic architecture with naive prompts (comparing monolithic to direct prompting yields larger gains than comparing neuro-symbolic to monolithic). However, the combination of both components achieved the highest performance and, critically, the greatest stability across different language models.

#### 3.2 Cross-Model Stability

Perhaps the most practically significant finding is the dramatic difference in cross-model stability between architectures. The neuro-symbolic system exhibited 10.8× lower variance in kappa values across five frontier language models compared to the monolithic alternative, with no model–measure combination falling below substantial agreement. This stability has important implications for system deployment and maintenance.

Healthcare AI systems must maintain consistent performance across model updates, vendor changes, and the continuous evolution of foundation models. A system whose performance varies from $\kappa = 0.020$ to $\kappa = 0.947$ depending on which language model backs it, as observed with direct prompting, cannot be reliably deployed in production. The neuro-symbolic architecture's stability (range: 0.710–0.947) reflects a fundamental property of the hybrid design: by constraining the language model to narrowly-scoped extraction tasks and delegating measure logic to symbolic rules, the architecture reduces dependence on any single model's idiosyncratic reasoning patterns.

This finding also has implications for regulatory approval and clinical validation. Systems demonstrating consistent performance across multiple model backends present a more tractable validation target than systems whose behavior is tightly coupled to a specific model version. As foundation models continue to evolve rapidly, architectures that isolate clinical logic from language understanding may prove more sustainable.

#### 3.3 Implications for Quality Measurement

The results suggest that automated abstraction could fundamentally alter the economics of quality measurement. Current estimates place the annual cost of quality reporting at $15.4 billion for U.S.

physician practices, with substantial portions allocated to manual chart abstraction [7]. If neuro-symbolic systems can reliably abstract measures at human-level accuracy, quality measurement could shift from sampling to census. Current practice often assesses quality on limited patient samples due to abstraction costs, constraining statistical power and delaying detection of quality problems. Automated abstraction at scale would enable continuous monitoring of every relevant case, transforming quality measurement from periodic assessment to real-time surveillance.

Reliable automation may also loosen the historical constraints that have shaped measure development. The requirement that concepts be reliably extractable by rules or human abstractors has filtered which measures reach implementation, biasing the quality measurement ecosystem toward simpler constructs. Measures requiring contextual interpretation, synthesis across report sections, or nuanced clinical judgment might become feasible if neuro-symbolic systems can operationalize them. The performance on CAP 43, a measure whose automated abstraction had previously been abandoned as too difficult, provides preliminary evidence for this possibility.

Beyond economics, automated abstraction could decouple quality monitoring from clinical workflow constraints. The COVID-19 pandemic revealed the fragility of measurement systems that compete with patient care for human attention; CMS granted reporting relief explicitly so organizations could redirect resources toward patients. Automated infrastructure would enable sustained measurement even during periods of healthcare system stress, ensuring that quality surveillance does not falter precisely when it matters most.

##### 3.4 Future Directions

Several extensions of this work merit investigation. The neuro-symbolic framework could be applied to quality measures beyond pathology, including measures requiring synthesis across multiple document types or longitudinal reasoning across encounters. The symbolic reasoning layer could incorporate clinical ontologies and knowledge graphs, potentially enabling automated detection of measure specification ambiguities or contradictions. The system's intermediate extraction outputs could also support quality improvement by identifying specific documentation patterns associated with measure failure, enabling targeted feedback to pathologists. On the methodological side, active learning approaches could reduce the development set size required for case-based refinement, lowering the barrier to deploying new measures. Prospective studies comparing automated versus manual abstraction in operational settings would provide definitive evidence of real-world utility.

#### 4 TRIPOD-LLM Reporting Checklist

This study is reported in accordance with the TRIPOD-LLM reporting guideline for studies using large language models [8]. Within the guideline's modular framework, this work falls under the *LLM methods* and *LLM evaluation in healthcare settings* research-design categories and the *classification* task category; items specific to de novo model development, instruction tuning, generative-text tasks, and summarization are therefore marked not applicable (N/A). In the table below, "Yes" indicates that an item is addressed, "Partial" that it is partially addressed, and "No" that it is not reported. Unless otherwise noted, locations refer to the main manuscript.

| Item | Checklist requirement (abbreviated) | Reported | Location / comment |
|---|---|---|---|
| 1 | Identify study as developing/evaluating an LLM; task, population, outcome | Partial | Title names the framework and clinical domain but does not explicitly state evaluation, target population, or outcome. |
| 2 | Structured abstract (per abstract checklist) | Yes | Structured abstract (Background, Methods, Results, Conclusions). |
| 3a | Healthcare context/use case and rationale, with references | Yes | Introduction. |
| 3b | Target population, intended use, and intended users | Partial | Introduction; intended use is stated, intended users only implicitly. |

*TRIPOD-LLM checklist (continued)*

| Item | Checklist requirement (abbreviated) | Reported | Location / comment |
|---|---|---|---|
| 4 | Study objectives; development/validation stage | Yes | Introduction (final paragraph) and Study Design. |
| 5a | Data sources per split and rationale | Yes | Methods (Dataset). |
| 5b | Data points; distribution; source, language, country | Partial | Methods (Dataset); counts and US source given, but language and detailed distribution not specified. |
| 5c | Dates of oldest and newest text items | Yes | Methods (Dataset): reports dated December 2023 to September 2025. |
| 5d | Data preprocessing and quality checking | Yes | Methods (Dataset): HL7 to FHIR mapping, manual completeness screening, de-identification (single source, so cross-institution comparison not applicable). |
| 5e | Handling of missing and imbalanced data | Partial | Reports screened for completeness; class imbalance addressed through prevalence-aware interpretation of $\kappa$ rather than resampling. |
| 6a | LLM name, version, and last training date | Yes | Supplement (LLM Inference Configuration); vendor training cutoffs not disclosed, so inference dates are reported instead. |
| 6b | Model architecture, training, and alignment | N/A | No de novo development or fine-tuning; hosted models used unchanged. System architecture is described in the Supplement. |
| 6c | Text-generation method; prompt engineering; inference settings | Yes | Methods (case-based refinement) and Supplement (LLM Inference Configuration): vendor-default, unseeded sampling. |
| 6d | Initial and postprocessed model output | Yes | Methods and Supplement: atomic LLM extractions combined by a symbolic layer into one of four determinations. |
| 6e | Classification details and rationale; thresholds | Partial | Four-class scheme defined (Methods); determinations come from a deterministic symbolic combiner, so no probability thresholds apply. |
| 7a | Quality metrics for generative outputs | N/A | Task is classification, not free-text generation. |
| 7b | Relevance of outcome metrics to the downstream task | Yes | Methods and Results: agreement (Cohen's $\kappa$) with the adjudicated gold standard. |
| 7c | Outcome definition; prediction method; inference date; metrics | Yes | Methods and Supplement; inference dates for the closed-weight models are reported. |
| 7d | Assessor qualifications, instructions, and agreement | Yes | Methods and Supplement (Abstractor Training); interrater reliability reported. Assessor demographics beyond professional background were not collected. |

*TRIPOD-LLM checklist (continued)*

| Item | Checklist requirement (abbreviated) | Reported | Location / comment |
|---|---|---|---|
| 7e | Comparison to other LLMs, humans, and benchmarks | Yes | Results: comparison with human abstractors, five LLM backends, and monolithic and direct baselines. |
| 8a | Labeling method; annotation guidelines with examples | Partial | Methods and Supplement describe the labeling scheme and CAP-based guidance; worked annotation examples are not included. |
| 8b | Number of annotators; proportion double-labeled; agreement | Yes | Two abstractors, 100% double-abstracted, interrater $\kappa$ reported (Methods and Results). |
| 8c | Annotator background and experience | Yes | Supplement (Abstractor Training). |
| 9a | Prompt design, curation, and selection | Yes | Methods and Supplement (case-based refinement). |
| 9b | Data used to develop prompts | Yes | Methods: prompts developed on the 400-report development set; evaluation set held out. |
| 10 | Preprocessing before summarization | N/A | Not a summarization task. |
| 11 | Instruction tuning and alignment details | N/A | No instruction tuning or alignment performed. |
| 12 | Compute or proxies (time, cost, inference time) | No | Not measured; acknowledged as a limitation in the Discussion. |
| 13 | Ethics committee approval; consent or waiver | Yes | The WCG Institutional Review Board determined the study exempt under 45 CFR 46.104(d)(4) (Protocol 20251925), a category that did not require informed consent (Methods). |
| 14a | Funding source and funder role | Yes | Funding section (main text). |
| 14b | Conflicts of interest and financial disclosures | Yes | Competing Interests section (main text). |
| 14c | Where the protocol can be accessed, or state none | Yes | A study protocol (version 1.0) was prepared and reviewed by the WCG Institutional Review Board (Protocol 20251925); it is available from the corresponding author on reasonable request. |
| 14d | Registration information, or state not registered | Yes | The study was not registered. |
| 14e | Availability of the study data | Yes | Data and Code Availability (main text): reports contain protected health information and are not publicly available. |
| 14f | Availability of code to reproduce results | Yes | Data and Code Availability (main text): code and prompts available from the corresponding author on reasonable request. |
| 15 | Patient and public involvement, or state none | Yes | No patient or public involvement. |
| 16a | Flow of data through study; counts with and without outcome | Yes | Methods, Results, and Figure 2: development and evaluation split with per-measure counts. |

*TRIPOD-LLM checklist (continued)*

| Item | Checklist requirement (abbreviated) | Reported | Location / comment |
|---|---|---|---|
| 16b | Characteristics per source and split; key dates; sample size | Partial | Sample sizes, splits, and report date range reported; per-source characteristics limited by the single-practice source. |
| 16c | Distribution of clinical variables, development vs evaluation | Partial | Outcome-label distributions differ by measure (discussed via $\kappa$ prevalence sensitivity); no formal covariate comparison across splits. |
| 16d | Number of participants and events per analysis | Yes | Methods and Results: per-measure and per-split counts and error counts. |
| 17 | Performance per prespecified metrics or human evaluation | Yes | Results: $\kappa$ values, comparisons, and confusion matrix. |
| 18 | Results of any LLM updating | N/A | No model updating performed. |
| 19a | Overall interpretation, including fairness | Partial | Discussion interprets results and compares with prior work; a formal fairness or subgroup analysis was not conducted. |
| 19b | Limitations; biases; uncertainty; generalizability | Yes | Discussion (Limitations). |
| 19c | Data challenges: representation, missingness, harmonization, bias | Partial | Single-source generalizability discussed; representation and missingness not analyzed in depth. |
| 19d | Intended input, end-user, and level of autonomy/oversight | Yes | Discussion: intended use (quality-measure abstraction for reporting, not patient care), input (full report text), intended users, and autonomous operation with auditable human oversight are specified. |
| 19e | Handling of poor-quality or unavailable input at deployment | Yes | Discussion: missing or ambiguous required elements are surfaced in the audit trace and can be routed for adjudication; operational handling of degraded inputs was not evaluated and is noted as a prerequisite for deployment. |
| 19f | Whether users interact with input or use; expertise required | Yes | Discussion: routine operation requires no per-report user interaction; introducing a new measure requires a clinician or clinical informaticist for case-based refinement and validation. |
| 19g | Next steps for future research; generalizability | Yes | Discussion (Future Directions), main text and Supplement. |

[3] Jacob Cohen. *Statistical Power Analysis for the Behavioral Sciences*. Lawrence Erlbaum Associates, Hillsdale, NJ, 2nd edition, 1988.
